# Understanding how a digital mental health intervention can be optimised to ensure effectiveness in the longer-term: findings from a causal mediation analyses of the CONEMO trials

**DOI:** 10.1101/2023.01.18.23284711

**Authors:** Nadine Seward, Wen Wei Loh, J. Jaime Miranda, Francisco Diez-Canseco, Heloisa Garcia Claro, Paulo Rossi Menezes, Ivan Filipe de Almeida Lopes Fernandes, Ricardo Araya

**Affiliations:** Centre for Global Mental Health, Health Service and Population Research Department, Institute of Psychiatry, Psychology & Neuroscience, King’s College London, UK. 18 De Crespigny Park, London, SE5 8AF, UK; Centre for Implementation Science, Health Service and Population Research Department, Institute of Psychiatry, Psychology & Neuroscience, King’s College London, UK. 18 De Crespigny Park, London, SE5 8AF, UK; Department of Data Analysis, Ghent University, Gent, Belgium. Henri Dunantlaan 1, 9000 Gent, Belgium; CRONICAS Centre of Excellence in Chronic Diseases, Universidad Peruana Cayetano Heredia, Lima, Peru, CRONICAS Centre of Excellence in Chronic Diseases, Universidad Peruana Cayetano Heredia, Av. Armendáriz 497, Miraflores, Lima 18, Peru; School of Medicine, Universidad Peruana Cayetano Heredia, Lima, Peru. Av. Honorio Delgado 430, San Martín de Porres 15102, Peru; The George Institute for Global Health, UNSW, Sydney, Australia. Level 5, 1 King Street Newtown NSW 2042 Australia; State University of Campinas. Cidade Universitária Zeferino Vaz - Barão Geraldo, Campinas - SP, 13083-970, Brazil; University of São Paulo. Rua da Reitoria 374 - Butantã São Paulo 05508-220 Sao Paulo Brazil

## Abstract

**Background:** Two CONEMO trials in Lima, Peru and São Paulo, Brazil evaluated a digital mental health intervention (DMHI) based on behavioural activation (BA) that demonstrated improvements in symptoms of depression between trial arms at three-months, but not at six-months. To understand how we can optimize CONEMO in the longer-term, we therefore aim to investigate mediators through which the DMHI improved symptoms of depression at six-months, separately for the two trials and then using a pooled dataset.

**Methods:** We used data that included adults with depression (Patient Health Questionnaire – 9 (PHQ-9) score ≥10) and comorbid hypertension and/or diabetes. Interventional effects were used to decompose the total effect of DMHI on symptoms of depression at six months into indirect effects via: understanding the content of the sessions without difficulty; number of activities completed that were self-selected to improve levels of BA; and levels of activation measured using the Behavioural Activation for Depression Short Form (BADS-SF).

**Findings:** Using the pooled dataset, understanding the content of the sessions without difficulty mediated a 10% [0.10: 95% CI: 0.03 to 0.15] improvement in PHQ-9 scores at six months; completing self-selected activities mediated a 12% improvement [0.12: 0.01 to 0.23]; and, lastly, BA mediated a 2% [0.02: 0.01, 0.05] improvement.

**Conclusions:** Our findings suggest that targeting participants to complete activities they find enjoyable will help to improve levels of activation and maintain the effect of the CONEMO intervention in the longer-term. Improving the content of the sessions to facilitate understanding can also help to maintain improvements.

## Introduction

Comorbidity of depression with other physical chronic conditions are on the rise, particularly in low- and middle-income countries (LMICs).(Moussavi *et al*., 2007) In Latin America, most of the disease burden is now explained by chronic diseases including depression, hypertension, and diabetes. Very often, depression amplifies the morbidity, and mortality associated with other chronic health conditions.(Gold *et al*., 2020) There is an urgent need for chronic disease management to include treatment for co-existing depression.(Lopez, Mathers, Ezzati, Jamison and Murray, 2006, Patel, 2000)

In addition to the above, there is also a large treatment gap for depression. In LMICs the proportion of people with severe mental health conditions who reported receiving any treatment was typically between 10-25%, compared to 50% or more in high-income countries.(Demyttenaere *et al*., 2004) One of the main reasons for this treatment gap is the shortage of specialised human resources and infrastructure such is the case in Latin American countries like Brazil and Peru.(Pan American Health Organization, 2018)

CONEMO (*Control Emocional* in Spanish and Portuguese) is a DMHI developed and tested in LMICs settings that aimed to integrate the management of depression with physical chronic conditions.(Araya *et al*., 2021) CONEMO is a six-week intervention based on behavioural activation for depression that took place in Lima, Peru, and São Paulo, Brazil. Although at three-months after enrolment, CONEMO was effective in improving PHQ-9 scores by at least 50% in the intervention arm compared to the enhanced usual care arm in both trials, there was no evidence to support this effect at six-months after baseline assessments.

Given the potential to bring this intervention to scale, it is important to understand how the CONEMO DMHI can be optimised to maximise benefits for this treatment in the long-term. The CONEMO trials conducted in Brazil and Peru are particularly insightful to address this, as the DMHI captured multiple forms of information on the sessions that could shed light as to why the effectiveness measured at three months did not hold at six months.(Araya *et al*., 2021)

The objective of our study is to use interventional indirect effects (Vansteelandt and Daniel, 2017) to investigate mediators that are causally intervening between the CONEMO DMHI and the improvement of depressive symptoms at six-months, including whether participants were able to understand the content of the sessions, the number of activities completed that were selected to improve levels of enjoyment and therefore activation, and levels of behavioural activation. We will investigate these mediators separately for the different sites, then using a pooled dataset.

## Methods

### Setting

This is a secondary data analysis using data from the CONEMO trials that took place between September 2016 and April 2018 in twenty health facilities in São Paulo, Brazil and seven in Lima, Peru.(Araya *et al*., 2021)

### Design

The trial in São Paulo was a cluster randomised trial whilst the trial in Lima was an individually randomised trial. Participants in both trials were eligible if they were aged 21, had hypertension and/or diabetes, and clinically significant depression symptoms (Patient Health Questionnaire-9 [PHQ-9] score ≥10).(Araya *et al*., 2021)

### CONEMO intervention

CONEMO is a low-intensity DMHI based on behavioural activation for depression and offered through a smartphone. CONEMO has 18 sessions delivered over a six-week period.(Araya *et al*., 2021) CONEMO was identical in both study sites, except it was supported by nurses in Lima and nurse assistants in São Paulo. Further details on the role of nurses can be found elsewhere.(Rocha, Aschar and Hidalgo-Padilla, 2021)

Participants in both study arms received enhanced usual care that consisted of being assessed for depression symptoms up to four times during the first month of the trial, and again during research follow-up assessments.(Araya *et al*., 2021)

To assess whether any short-term (three-months) improvements were maintained over time, all participants in both arms were assessed again at six-months. Details of the main trial methodology and results can be found elsewhere.(Araya *et al*., 2021)

### Measures

#### Exposure

Participants in the experimental arm were offered the CONEMO DMHI (exposed) whereas participants in the control arm were offered enhanced usual care (unexposed).

#### Outcome

The trials used the primary outcome of an improvement of at least 50% in PHQ-9 scores between baseline and three-month assessments. For our mediation analysis, we used the outcome of improvement of at least 50% in PHQ-9 scores between baseline and the six-month follow-up visit. In doing so, we can establish temporality between the mediators and the outcome to rule out reverse causation, as our mediators were measured earlier at the three-month assessments.

#### Mediators

A mediator is defined as an intervening variable on a causal pathway between an exposure and an outcome.(Lee *et al*., 2021) Mediators measured only in the experimental arm are variables that are a direct consequence of receiving the DMHI and therefore not measured for participants in the control arm. Mediators were included in the final model if they were associated with another mediator, or the outcome whilst adjusting for mediator-outcome confounders (*p*<0.10).

Mediators measured in both arms of the trial were only included in the final model if they were associated with the CONEMO intervention or the outcome (*p*<0.10) whist adjusting for mediator-outcome confounders. Using the above criteria for our two types of mediators, we can capture any potential mediator that is influenced by the intervention and influences either another mediator, or the improvement in depressive symptoms.

Causal meditation analysis requires measuring levels of mediators for participants who were exposed, as well as the unexposed (i.e. counterfactual). Values of mediators measured in both arms of the trials are set as unexposed for participants in the control arm and exposed for participants in the experimental arm. The values of mediators measured in the experimental were discretized so that an “unexposed” value of zero was defined for those who were assigned to the experimental arm but were not exposed to the mediator of interest (i.e. did not complete any of the assigned activities). After adjusting for relevant confounders, we assumed that participants in the control arm who could not have accessed the DMHI, had the same unexposed value of zero as participants in the experimental arm who were not exposed to the mediator of interest.

Based on the above criteria, the following mediators were selected:

##### The messages on the DMHI are understood without difficulty (M1 – measured in the experimental arm only at the three-month follow-up visit)

Whether or not a participant understands the content of the messages delivered through the DMHI can influence not only whether a participant completes activities as intended, but also levels of behavioural activation measured at three-months. To capture this effect, we included a variable that ascertained whether the participants understood the content of the sessions delivered via the DMHI without difficulty (M1=no-unexposed/yes - exposed) and was captured in the experimental arm only in the assessment conducted three-months after baseline.

##### Number of assigned activities on the DMHI completed (M2 – measured in the experimental arm only and captured automatically via the DMHI when participants completed a session)

At each session, the DMHI relayed messages that encouraged participants to select and complete a set of activities they found enjoyable. M2 captures the number of activities that a participant reports completing, and it was categorised into the following: no assigned activities completed (unexposed); 1-10 assigned activities completed; and 11-27 assigned activities completed.

##### Level of behavioural activation (M3 – measured in both trial arms at three months)

Behavioural activation was measured for participants in both the intervention (exposed) and control arms (unexposed) at three-months from baseline. The Behavioural Activation for Depression Scale Short Form (BADS-SF) was used to assess the level of behavioural activation achieved with the CONEMO DMHI and was measured in both experimental (exposed) and control arms (unexposed).(Manos, Kanter and Luo, 2011) BADS-SF scores range from zero to 54, with higher scores indicating higher levels of behavioural activation.

##### Dependency of mediators on one another

A by-product of the interventional indirect effects is that in addition to the mediator-specific indirect effects, there is another indirect effect via the mediators’ mutual dependence on one another.(Vansteelandt and Daniel, 2017) This indirect effect is close to zero when mediators are conditionally independent and non-zero when mediators interact or covary in their effects on the outcome.

##### Mediator-outcome confounders

Due to the randomised nature of the exposure, it is not necessary to account for confounders of the association between the exposure and the outcome. However, it is necessary to account for baseline confounders (unaffected by the intervention) that were associated with the mediator and the outcome. These mediator-outcome confounders can generate spurious correlations between the mediator and outcome when unadjusted for, potentially distorting these associations. We considered more than thirty potential baseline characteristics that are not influenced by the intervention, as potential confounders. The selection process for these confounders is described in the section on *estimation methods* below.

### Statistical analysis

#### General

To better understand the relationship between different mediators and improved symptoms of depression, we compare different mediators and confounders with the outcome (a PHQ-9 score reduction of at least 50% between baseline and six-months) separately for the different trials. Differences in baseline characteristics between treatment arms can be found in the main trial publication.(Araya *et al*., 2021)

#### Mediation analysis

We aimed to investigate the extent to which improved symptoms of depression at six-months after baseline, captured using the PHQ-9 questionnaire, was explained via selected mediators. To achieve this, we used the interventional indirect effects approach for causal mediation analysis to understand population level effects relevant to this analysis.(Vansteelandt and Daniel, 2017) We applied this approach to mediation separately for the different CONEMO trials, then using the pooled dataset. The final model that was considered for this mediation analysis in both trials is shown in Figure 1. The mediation analysis conformed to AGReMA statement for reporting mediation analysis of randomised trials.(Lee *et al*., 2021)

**Figure 1:**
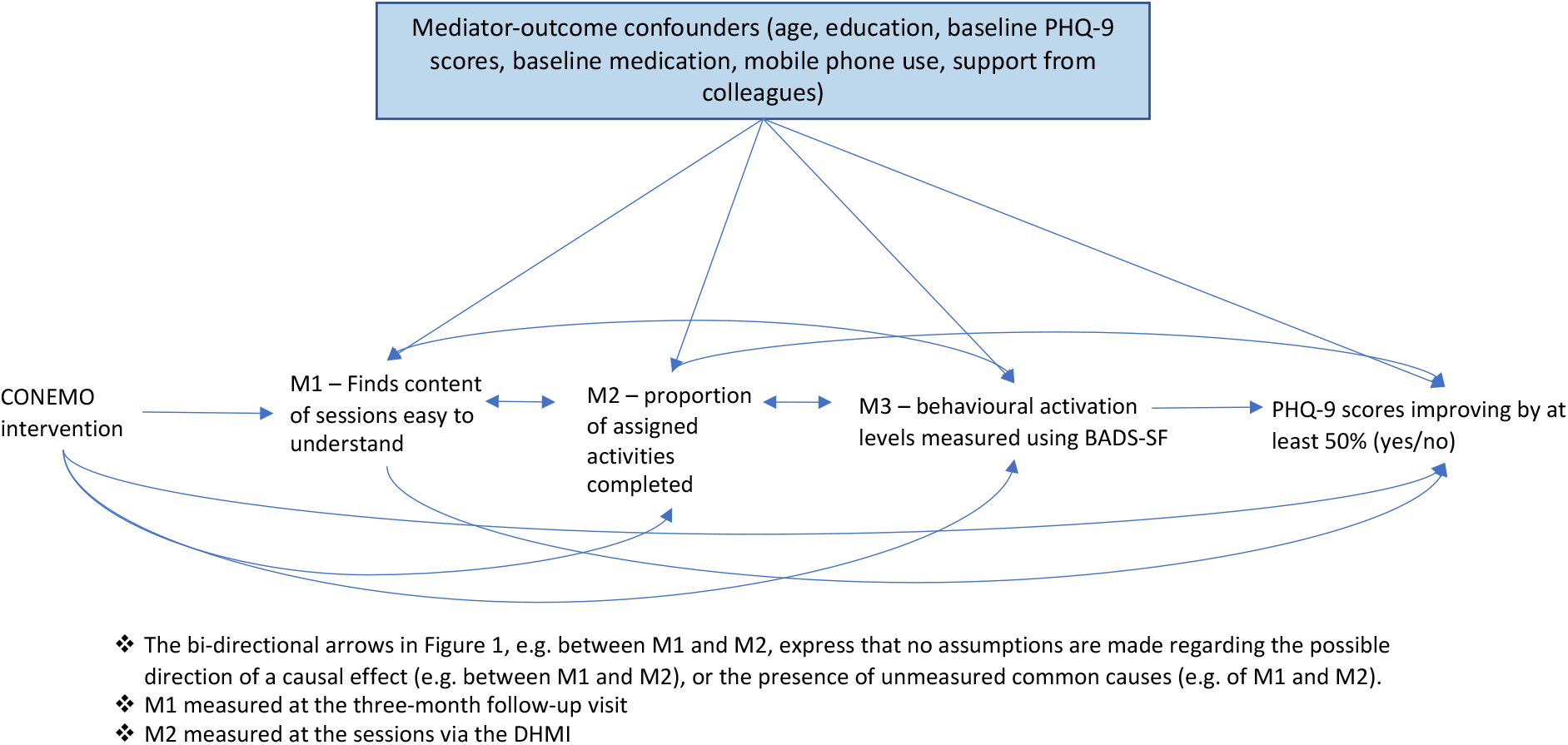
Causal model demonstrating the proposed mediating pathways through which the CONEMO intervention may improve PHQ-9 scores in Lima and São Paulo at the six-month follow-up visit

##### Decomposition of total effect of the CONEMO intervention into direct and indirect effects

We decomposed the total effect of the CONEMO intervention into interventional indirect effects via each of the three posited mediators and the direct effect via none of the mediators. The interventional indirect effect via a particular mediator (e.g., understanding content of the sessions without difficulty) can be interpreted as the average change in the potential outcome (improved symptoms of depression at six months) resulting from shifting the counterfactual distribution of that mediator from the exposed status (e.g., understand content of the sessions without difficulty) to the unexposed status (found content of sessions difficult to understanding), while setting each of the remaining mediators to random draws from either the exposed or unexposed group, depending on the specific decomposition. In doing so, valid inferences are not contingent on strict assumptions, such as correctly specifying the causal ordering among the mediators which are unlikely to be feasible in our current setting and can lead to incorrect inferences when violated. (Loh, Moerkerke, Loeys and Vansteelandt, 2020)

##### Estimation and model fit

Estimation for the interventional indirect effects was based on Monte Carlo integration using a 1,000-fold expanded dataset. (15) The expanded dataset was created in four steps separately for each of the different sites. In the first step, we fitted a model for each mediator given exposure and other predictors. Specifically, we fitted logistic (M1), ordinal (M2), and linear (M3) regression models to the observed data. Each model included a combination of predictors that were shown to be associated with the mediator of interest at the 10% level including: age, education, baseline PHQ-9 scores, baseline medication to treat a psychological problem, support from work colleagues, and use of a mobile phone. Interactions and non-linearities were explored and included if determined to be significant at the 10% level, using Stata’s post estimation command, *testparm*. The models for the pooled analyses also included a dummy variable representing the study site where interactions between study sites, and the different predictors were explored using similar criterion described above.

In the second step, the fitted mediator models were used to generate random, subject-specific Monte Carlo draws of each mediator for both the exposed and unexposed condition (i.e. counterfactual), given their observed covariate values.

In the third step, we fitted a model for the outcome, using a logistic regression model for recovery from depression, separately in the exposed and unexposed, given the mediators and mediator-outcome confounders. Any potential mediator-outcome confounder was included if it was associated with the mediator or the outcome (*p*<0.10). Models for the pooled dataset included and mediator-outcome confounder included in either of the individual sites as well as a variable to represent study site. We used a model selection criterion similar to that of the mediator models; i.e., Any relevant non-linearities and interactions were included in the outcome model if determined to be significant at the ten percent level, using the post-estimation *testparm* command in Stata.

Mediator-outcome confounders included in the São Paulo analyses include baseline medication to treat a psychological problem (yes/no), age (continuous), baseline PHQ-9 scores, number of years in education, extent to which a mobile phone is used in everyday activities (i.e. sending receiving messages, video calls, booking appointments, taking pictures, playing games, etc). Confounders in the Lima analyses were similar except for the addition of having social support from a friend or colleague (this was represented by number of times they would talk to a friend or colleague in a month).

The São Paulo dataset had the following interactions: BAD-SF scores measured at three-months (M3) and medication taken to treat a psychological problem measured at baseline; and whether participants found the content of the sessions difficult to understand (M1) and baseline PHQ-9 scores. The Lima dataset had an interaction between assigned activities completed (M2) and baseline BAD-SF scores. Continuous variables (BAD-SF at baseline and three-months (M3) and PHQ-9 at baseline) that were included in an interaction term were mean centred to reduce multicollinearity.

The pooled dataset included any mediator-outcome confounder used for the separate study sites, as well as a dummy variable representing study sites. Interactions between the dummy variable, mediators and mediator outcome confounders were also examined for and included if the above-described criterion were fulfilled. Significant interactions were found between the following: study site and baseline medication; study site and baseline PHQ-9 scores, BAD-SF at three-months (M3) and study site; BAD-SF at three-months and medication; and understanding content of sessions without difficulty (M2) and baseline PHQ-9 score.

In the fourth step, we used the fitted outcome model to predict the potential outcomes in the expanded dataset given the random, subject-specific draws of the mediator counterfactuals from the second step. The interventional indirect effects were then calculated as the average differences between potential outcomes under different hypothetical exposure levels.

Bias-corrected confidence intervals were based on nonparametric bootstrap with 1,000 resamples that adjusted for clustering and stratification.(Vansteelandt and Daniel, 2017)

#### Assumptions

Due to the randomised nature of the CONEMO trials, the assumptions about no unmeasured confounders between the exposure and each mediator, and between the exposure and the outcome, are fulfilled. The main assumption relevant to our study is that there is no unmeasured -mediator-outcome confounders.

#### Missing data

In São Paulo out of the 880 participants enrolled at baseline, 656 (85%) had data available for the mediation analyses. In Lima, out of 432 participants enrolled at baseline, 389 (90%) had data available for the mediation analysis. Due to the non-random nature of the missing data, we were not able implement the imputation models (Supplement 1).

#### Sensitivity analyses

Primary outcomes for depression are mainly reported as either recovery from depression (PHQ-9 <10) or an improvement of at least 50% in PHQ-9 scores between baseline and follow-up. We selected our outcome measure of improved symptoms of depression (50% reduction in PHQ-9 scores between baseline and the six month follow-up) to ensure consistency with the outcomes reported in the main trial.(Araya *et al*., 2021) To facilitate comparability with other studies, we also conducted the same analyses using recovery from depression at six-months (PHQ-9 <10).

### Ethical approval and consent

In Brazil, the trial protocol received ethical approval by the Institutional Review Board at the University of São Paulo (nº 457.605) and the National Commission of Ethics in Research (nº 355.039). In Lima, the protocol was approved by the Institutional Review Board at the Universidad Peruana Cayetano Heredia (nº 34516-16).

Role of funding source: Only the original CONEMO trials received funding and they and for this, they had no role in this study design, collection, analysis, and interpretation of data. Nor did the funders have any involvement in this mediation analyses, including for instance the writing of the report, and in the decision to submit the paper for publication.

## Results

### General

Table 1 compares mediators in the experimental arm only, between participants with improved PHQ-9 scores of least 50% between baseline and six-months, and those with scores that did not improve by this amount. Without adjustment for mediator-outcome confounders, descriptive statistics show that participants who had improved symptoms of depression at six-months after baseline were more likely to have understood the content of the sessions without difficulty (M1) in both Lima and São Paulo. In both sites, participants who completed their assigned activities on the DMHI, were also more likely to have improved symptoms of depression at six-months (M2). Lastly, findings in Lima and São Paulo suggest that at six-months, participants who showed improvement in depression symptoms, had higher mean behavioural activation levels (M3).

**Table 1.**
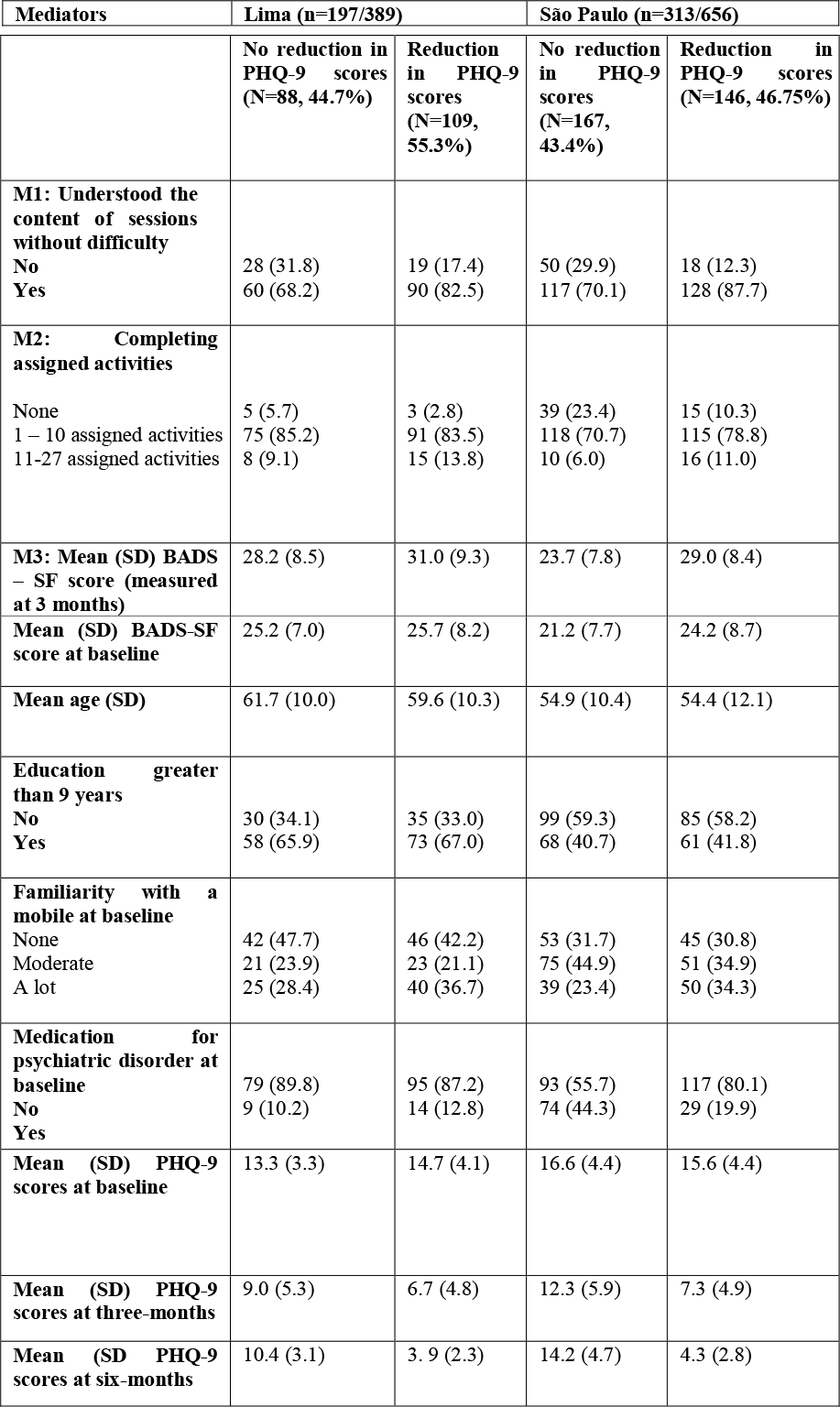
Comparison of mediators between participants with improved symptoms of depression (a reduction in PHQ-9 scores of at least 50% at six-month from PHQ-9 scores at baseline) in the experimental arm of the trial only

### Mediation analyses

Results demonstrated that at six-months after baseline, there was some evidence to support a small difference in the probability of symptoms of depression improving by at least 50% (yes/no) between the experimental and control arm using data from the pooled dataset (0.04 [95% bias-correct confidence interval: 0.00 to 0.09]), but not in the separate trials in Lima and São Paulo (Table 2).

**Table 2:**
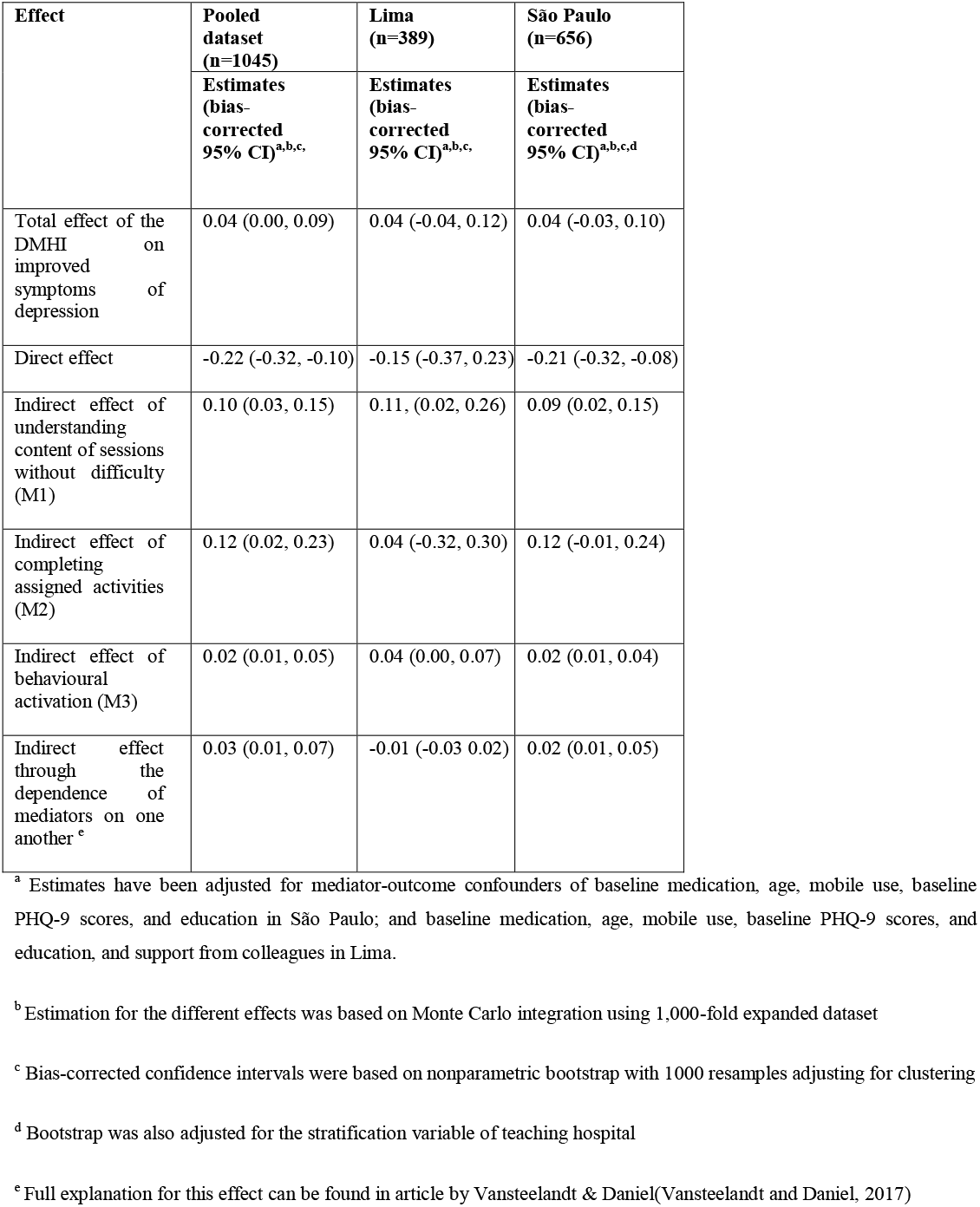
Total effect and interventional (in)direct effects of the CONEMO intervention on improved symptoms of depressions measured depression (a reduction in PHQ-9 scores of at least 50% at six-month from PHQ-9 scores at baseline) at six-months follow-up

Finding the content of the sessions easy to understand (M1) mediated a 10% difference in the probability of symptoms of depression improving by at least 50% (yes/no) in the pooled dataset (difference in probability of symptoms of depression improving by at least 50% between participants who understood the content of the sessions without difficulty, compared to those who found the content of the sessions difficult to understand: 0.10 [95% bias-corrected confidence interval: 0.03 to 0.15]), as well as in Lima (0.11 [0.02 to 0.26]) and São Paulo (0.09 [0.02 to 0.15]) separately.

Estimates using data from the pooled dataset provide evidence that completing the activities as intended (M2) was responsible for mediating a 12% difference in symptoms of depression improving by at least 50% (mean difference in symptoms of depression improving by at least 50% between participants who completed assigned activities compared to those who did not complete any assigned activities: 0.12 [0.02 to 0.23]). There was some evidence to support this finding in São Paulo (0.12 [-0.01 to 0.24), however there was no evidence to support the same finding in Lima (0.04 [-0.32 to 0.30]).

There was evidence that behavioural activation (M3) mediated a 2% difference in symptoms of depression improving by at least 50% using the pooled dataset (mean difference in proportion with improved symptoms of depression between participants with BAD-SF scores in the exposed population, compared to participants with BAD-SF scores in the unexposed population: 0.02 [95% bias-corrected confidence interval: 0.01 to 0.05]), as well as in São Paulo (0.02 [0.01 to 0.04]), and a 4% difference in Lima (0.04 [0.00, 0.07]) respectively.

Lastly, using the pooled dataset, the indirect effect attributable to the mutual dependence of the mediators on one another, was a 3% difference in symptoms of depression improving (0.03 [95% confidence interval: 0.01 to 0.07]), and a 2 %, difference in São Paulo (0.02 [0.01, 0.05] but there was no evidence to support this in Lima (−0.01 [-0.03, 0.02]).

### Sensitivity analyses

Estimates from our analyses using as outcome the probability of recovery from depression between the experimental and control arm at six-months (defined as PHQ-9 score <10 at the sixth month follow-up visit), were largely consistent with the findings using the outcome measure of a reduction in PHQ-9 scores of at least 50% between baseline and the sixth month follow-up. Findings are reported in Supplement 2.

## Discussion

This novel application of the causal mediation-based framework of interventional indirect effects provides insight into how the CONEMO DMHI can be adapted to achieve better longer-term improvements in depressive symptoms. Whilst simultaneously accounting for all mediators, we have been able to successfully demonstrate the importance of ensuring the content of the app sessions is understood without difficulty. Importantly, the indirect effects of the number of assigned activities completed that were selected to improve levels of activation (M2) and activation levels captured using the BADS-SF (M3), confirm the importance of targeting behavioural activation to improve depression outcomes in the longer term. Our findings, benefit from the evaluation of the same DMHI in two different contexts, thus allowing for the assessment of how mediators operate in diverse real-world settings.

A priori, we theorised that understanding session content without difficulty (M1) will influence the number of activities completed that were selected to improve behavioural activation (M2), and therefore activation levels (M3). The first two mediators are necessary pre-conditions required to maximise the benefits of the DMHI, that was based on behavioural activation for depression. This theory was supported by our findings that indicated the mutual dependence of the mediators on one another, mediated an improvement in our outcome.

Findings from both study sites and the pooled analyses suggest that the intervention was less effective at six-months among those participants who found the content of the sessions difficult to understand. Adapting the DMHI so that the sessions are understood without difficulty by all participants, can also help to ensure that more activities are completed, and levels of behavioural activation are higher. Our findings that both the number of activities completed (M2) as well as activation levels (M3) mediated improvements in depression outcomes, confirm the importance of psychological interventions targeting behavioural activation. It is important to note that these two later mediators capture different properties of behavioural activation. Whereas the number of activities completed, is an indicator as to the dose-response of the intervention that targets activation, the BADS-SF captures levels of activation after the activities were completed as measured by the questionnaire that also includes items unrelated to activity levels

Findings from this mediation analysis are also supported by qualitative research arising for the CONEMO trial in Lima that explored participants and nurses’ experiences in using the DMHI. Some recommendations include making the DMHI more user-friendly by writing session content in simpler text and using more videos. These adaptations could potentially reduce the role of nurses employed on a full-time basis in Lima, that is not feasible if brought to scale.(Toyama, 2022)

There has been conflicting evidence to support the role of behavioural activation in mediating the effects of psychological therapies in sessions that are delivered both face to face and through digital mental health interventions.(Domhardt *et al*., 2021, Lemmens, 2016) However, many of the studies are heterogeneous with differences in the content of sessions, levels of depression, taking place in diverse contexts, often coupled with poor methodological quality making comparisons difficult.(Domhardt *et al*., 2021, Janssen *et al*., 2021, Nasrin, Rimes, Reinecke, Rinck and Barnhofer, 2017, Richards *et al*., 2017) More recently, a mediation analysis using data from a trial in Goa, India, using the same approach as in this paper, demonstrated a significant role of behavioural activation in mediating a reduction in symptoms of depression, but not attending sessions or completing assigned activities.(Patel *et al*., 2017, Seward, Vansteelandt, Moreno-Agostino, Patel and Araya, 2022) Ensuring future mediation analyses use robust approaches to mediation, as suggested by the AGReMA statement, will help to better understand the mechanisms through which psychological interventions improve outcomes of depression.(Lee *et al*., 2021)

Our approach to meditation has several strengths. Importantly, our mediation analyses allowed us to include multiple mediators, their interactions and non-linearities. Failing to simultaneously account for the different mediating pathways by excluding mediators could over or underestimate the indirect and direct effects. Our analyses also had a vast array of data that was collected via the DMHI tool that provided information that allowed us to estimate the effects in a less biased approach than having a participant recall at each session whether activities were completed or not.

When applying the interventional effects to a randomised trial, the main underlying assumption is that all important mediator-outcome confounders are accounted for. Failing to do so, can also potentially bias all estimates including the direct and indirect effects. Another issue is setting the mediators to a random subject-specific draw when there are high dimensional data (multiple predictors and interactions) can lead to bias in correctly specifying the model. One of the more novel methods to mediation analysis uses a combination of machine learning and interventional effects that can overcome this limitation.(Benkeser and Ran, 2021) These analyses benefit from pooled data from two trials testing the same intervention and uses a robust approach to causal mediation analysis to understand how a complex psychological therapy delivered using a DMHI, improved symptoms of depression.

The future of mHealth applications, facilitated by the gradual transition toward remote care in recent years arising during the COVID-19 pandemic, shows great promise to reduce existing large treatment gaps in mental health, especially with the rapid development of robust methodologies to understand how to adapt intervention to improve outcomes, including causal machine learning(Benkeser and Ran, 2021) and robust implementation research.(Seward *et al*., 2021)

## Supporting information

Supplement file 1 missing data

Supplement file 2 sensitivity analyses

## Data Availability

Data will be made available publicly once all analyses from trial data is complete

## Required statements

## Acknowledgments

We would like to thank the authors and all of those involved in the original CONEMO trials including the trial participants. We would also like to acknowledge the funders of the original trial, the National Institute of Mental Health (U19MH098780).

## Contributors

NS and RA conceptualised the study. NS and completed the main analyses and WWL advised. All authors were involved with the overall design of the mediation analyses. NS wrote the original draft and all authors critically revised and approved the manuscript. NS, RA, had full access to all the data in the study. All authors had final responsibility for the decision to submit for publication.

## Data sharing

Data will be made available once the papers associated with this trial have been published.

