## Supplement file 1 missing data for "Understanding how a digital mental health intervention can be optimised to ensure effectiveness in the longer-term: findings from a causal mediation analyses of the CONEMO trials"

**Supplement 2: Differences in missing data between participants who are exposed to a mediator and participants who are unexposed in Lima and Sao Paulo**

Indirect effects are estimated by calculating the difference in the probability in symptoms of depression improving by at least 50%(yes/no) when changing levels of mediator between exposed and unexposed status, for participants in the experimental arm. Therefore, any missing data that influences this difference, could bias the indirect effects estimated using interventional effects.

For the first two mediators this difference is calculated by subtracting probability of outcome occurring for participants in the experimental arm who are unexposed (found sessions difficult to understand (M1) and did not complete any activities (M2)) from probability of outcome occurring in the exposed (understood content of sessions (M1) and completed assigned activities (M2). For M3, this difference is calculated by subtracting probability of outcome occurring for participants in the experimental arm with levels of M3 in control arm, from probability of outcome occurring for participants in experimental arm with levels of M3 set to levels participants experience in experimenting arm.

Therefore, for M1 and M2, missing data for both the mediator in question, or the outcome measure could bias estimates. As an example, participants who did not complete any activities were more likely to drop out of the trial and therefore recover from depression. This could underestimate the mediating effects of completing assigned activities self selected to improve activation levels to recover from depression.

**Lima: Table demonstrating association between missing information on mediators and the outcome of a reduction in symptoms of depression by at least 50%**

| **Mediators** | **Control arm (n=215)** | | | **Experimental arm (n=217)** | | |
| --- | --- | --- | --- | --- | --- | --- |
|  | **Symptoms did not improve**  **(N=93, 43.2%)** | **Symptoms improved**  **(N=99, 46.0%)** | **Missing**  **(N=23, 10.7%)** | **Symptoms did not improve (N=88, 40.6%)** | **Symptoms improved (N=109, 50.2%)** | **Missing**  **(n=20, 9%)** |
| **M1: Understood the content of sessions without difficulty**  **No**  **Yes**  **Missing** | .. | .. | .. | 28 (31.8)  60 (68.2) | 19 (17.4)  90 (82.6) | 1 (16.7)  6 (83.3) |
| **M2: Completing assigned activities**  None  1 – 10 assigned activities  11-27 assigned activities  Missing | .. | .. | .. | 7 (7.7)  76 (83.5)  8 (8.8) | 4 (3.6)  93 (83.0)  15 (13.4) | 8 (40.0)  10 (50.0)  2 (10.0) |
| **M3: Mean (SD) BADS – SF score (measured at 3 months)**  **Missing** | 24.0 (8.7) | 27.7 (8.9) | 26.9 (11.3) | 28.4 (8.4) | 31.0 (9.2) | 25.0 (10.5) |

**São Paulo: Table demonstrating association between missing information on mediators and the outcome of a reduction in symptoms of depression by at least 50%**

| **Mediators** | **Control arm (n=440)** | | | | **Experimental arm (n=440)** | | |  |
| --- | --- | --- | --- | --- | --- | --- | --- | --- |
|  | **Symptoms did not improve**  **(n=226, 51.4%)** | **Symptoms improved**  **(n=153, 34.8%)** | **Missing**  **(n=94 13.9%)** | **Total (n=440, 100%)** | **Symptoms did not improve (n=202, 45.9%)** | **Symptoms improved (n=173, 39.3%)** | **Missing**  **(n=65, 14.8%)** | **Total (n=440, 100%)** |
| **M1: Understood the content of sessions without difficulty**  **No**  **Yes**  **Missing** | .. | .. | .. |  | 55 (68.8)  19 (12.75)  6 (7.5) | 119 (45.6)  132 (50.6)  10 (3.8) | 28 (28.3)  22 (22.2)  49 (49.5) | 202 (45.9)  173 (39.3)  65 (14.8) |
| **M2: Completing assigned activities**  None  1 – 10 assigned activities  11-27 assigned activities  Missing | .. | .. | .. |  | 66 (32.7)  88 (43.6)  48 (23.8)  0 | 35 (20.2)  82 (47.4)  56 (32.4)  0 | 48 (73.9)  12 (18.5)  5 (7.7)  0 | 149 (33.9)  182 (41.4)  109 (24.8)  0 |
| **M3: Mean (SD) BADS – SF score (measured at 3 months)**  **Missing** | 23.8 (8.4) | 26.8 (8.8) | 25.6(8.6) | 27 (8.6) | 23.6 (7.9) | 28.8 (8.2) | 25.4 (7.3) | 26.0 (8.4) |
