## Supplement file 2 sensitivity analyses for "Understanding how a digital mental health intervention can be optimised to ensure effectiveness in the longer-term: findings from a causal mediation analyses of the CONEMO trials"

**Supplement 3: Sensitivity analyses comparing estimates from interventional indirect effects between the outcome of recovery from depression at six-months and reduction in PHQ-9 scores by 50% between baseline and six-months in the pooled analyses, the Lima trial, and the trial in São Paulo**

**Table 1: Pooled analysis (n=1045)**

| **Effect** | **Reduction in PHQ-9 scores by 50% between baseline and sixth months** | **Recovery from depression at six months (PHQ-9 scores less than 10)** |
| --- | --- | --- |
|  | **Estimates (bias-corrected 95% CI)^a,b,c^** | **Estimates (bias-corrected 95% CI)^a,b,c^** |
| Total effect on recovery from depression | 0.04 (0.00, 0.09) | 0.05 (0.00, 0.10) |
| Interventional direct effect | -0.22 (-0.32, -0.10) | -0.18 (-0.29, -0.08) |
| Understanding content of sessions without difficulty (M1) | 0.10 (0.03, 0.15) | 0.08 (0.02, 0.16) |
| Completing assigned activities (M2) | 0.12 (0.02, 0.23) | 0.11 (0.00, 0.20) |
| Levels of behavioural activation (M3) | 0.02 (0.01, 0.05) | 0.02 (0.01, 0.05) |
| Interventional indirect effect through the dependence of BA levels on sessions (M3) | 0.03 (0.01, 0.07) | 0.02 (0.01, 0.06) |

^a^ Estimates have been adjusted for mediator-outcome confounders of baseline PHQ-9 scores, use of mobile phone, education, age, baseline medication to treat depression, and support from colleagues

^b^ Estimation for the different effects was based on Monte Carlo integration using 1,000-fold expanded dataset

^c^ Bias-corrected confidence intervals were based on nonparametric bootstrap with 1000 resamples adjusting for clustering

**Table 2: Lima (n=389)**

| **Effect** | **Reduction in PHQ-9 scores by 50% between baseline and sixth months** | **Recovery from depression at six months (PHQ-9 scores less than 10)** |
| --- | --- | --- |
|  | **Estimates (bias-corrected 95% CI)^a,b,c^** | **Estimates (bias-corrected 95% CI)^a,b,c^** |
| Total effect on recovery from depression | 0.04 (-0.03, 0.12) | 0.09 (0.02, 0.16) |
| Interventional direct effect | -0.14 (-0.37, 0.19) | -0.09 (-0.43, -0.18) |
| Understanding content of sessions without difficulty (M1) | 0.11, (0.03, 0.26) | 0.05 (-0.07, 0.18) |
| Completing assigned activities (M2) | 0.04 (-0.29, 0.29) | 0.11 (-0.16, 0.39) |
| Levels of behavioural activation (M3) | 0.05 (0.01, 0.10) | 0.02 (-0.01, 0.05) |
| Interventional indirect effect through the dependence of BA levels on sessions (M3) | -0.02 (-0.05, 0.01) | -0.01 (-0.03, 0.02) |

^a^ Estimates have been adjusted for mediator-outcome confounders of baseline PHQ-9 scores, use of mobile phone, education, age, baseline medication to treat depression, and support from colleagues

^b^ Estimation for the different effects was based on Monte Carlo integration using 1,000-fold expanded dataset

^c^ Bias-corrected confidence intervals were based on nonparametric bootstrap with 1000 resamples adjusting for clustering

**Table 2: São Paulo (n=656)**

| **Effect** | **Reduction in PHQ-9 scores by 50% between baseline and sixth months** | **Recovery from depression at six months (PHQ-9 scores less than 10)** |
| --- | --- | --- |
|  | **Estimates (bias-corrected 95% CI)^a,b,c^** | **Estimates (bias-corrected 95% CI)^a,b,c^** |
| Total effect on recovery from depression | 0.05 (-0.03, 0.11) | 0.03 (-0.05, 0.08) |
| Interventional direct effect | -0.22 (-0.37, -0.03) | -0.23 (-0.34, -0.12) |
| Understanding content of sessions without difficulty (M1) | 0.13 (0.05, 0.22) | 0.13 (0.05, 0.18) |
| Completing assigned activities (M2) | 0.13 (-0.01, 0.30) | 0.09 (-0.01, 0.18) |
| Levels of behavioural activation (M3) | 0.01 (-0.04, 0.04) | 0.01 (-0.03, 0.03) |
| Dependency of mediators on one another (M3) | -0.02 (-0.08, 0.08) | 0.03 (0.02, 0.06) |

^a^ Estimates have been adjusted for mediator-outcome confounders of baseline medication, age, mobile use, baseline PHQ-9 scores, and education

^b^ Estimation for the different effects was based on Monte Carlo integration using 1,000 fold expanded dataset

^c^ Bias-corrected confidence intervals were based on nonparametric bootstrap with 1000 resamples adjusting for clustering and stratification of teaching hospital

**_**
